# Aerosol measurement identifies SARS-CoV 2 PCR positive adults compared with healthy controls

**DOI:** 10.1101/2022.01.21.22269423

**Authors:** Desireé Gutmann, Gerhard Scheuch, Timon Lehmkühler, Laura-Sabine Herrlich, Martin Hutter, Christoph Stephan, Maria Vehreschild, Yascha Khodamoradi, Ann-Kathrin Gossmann, Florian King, Frederik Weis, Maximilian Weiss, Holger F Rabenau, Juergen Graf, Helena Donath, Ralf Schubert, Stefan Zielen

## Abstract

**Background:** SARS-CoV-2 is spread primarily through droplets and aerosols. Exhaled aerosols are generated in the lung periphery by ‘reopening of collapsed airways’. Aerosol measuring may detect highly contagious individuals (“super spreaders or super-emitters”) and discriminate between SARS-CoV-2 infected and non-infected individuals. This is the first study comparing exhaled aerosols in SARS-CoV-2 infected individuals and healthy controls.

**Design:** A prospective observational cohort study in 288 adults, comprising 64 patients testing positive by SARS CoV-2 PCR before enrollment, and 224 healthy adults testing negative (matched control sample) at the University Hospital Frankfurt, Germany, from February to June 2021. Study objective was to evaluate the concentration of exhaled aerosols during physiologic breathing in SARS-CoV-2 PCR-positive and -negative subjects. Secondary outcome measures included correlation of aerosol concentration to SARS-CoV-2 PCR results, change in aerosol concentration due to confounders, and correlation between clinical symptoms and aerosol.

**Results:** There was a highly significant difference in respiratory aerosol concentrations between SARS-CoV-2 PCR-positive (median 1490.5/L) and -negative subjects (median 252.0/L; p<0.0001). There were no significant differences due to age, sex, smoking status, or body mass index. ROC analysis showed an AUC of 0.8918.

**Conclusions:** Measurements of respiratory aerosols were significantly elevated in SARS-CoV-2 positive individuals and may become a helpful tool in detecting highly infectious individuals via a noninvasive breath test.

**Clinical Trial Number:** ClinicalTrials.gov Identifier: NCT04739020.

**Summary of the main point:** In this prospective, comparative cohort study, higher numbers of exhaled respiratory aerosols correlate with a positive PCR test for SARS-CoV-2. Measurement of exhaled aerosols may become a helpful tool in detecting contagious individuals via a readily available breath test.

## Introduction

By July 2021, the severe acute respiratory syndrome coronavirus 2 (SARS-CoV-2) pandemic had caused more than 187 million confirmed cases and four million deaths.^1–3^ As the prevalence of SARS-CoV-2 infection and associated pulmonary disease (coronavirus disease–2019 [COVID-19]) remain high across the globe, the pandemic has been one of the greatest threats to the global economy and social infrastructure.^4^ Current research suggests that SARS-CoV-2 is spread primarily through droplets and aerosols.^5,6^ In addition to symptomatic carriers, asymptomatic infections and highly contagious carriers (‘super spreaders’) are key drivers of virus spread.^7–10^

Aerosols are defined as a suspension of solid or liquid particles within a gas mixture (e.g., air); ^11–13^ while droplets are defined as particles of a size approximately >100 µm. During normal breathing, small aerosol particles can be detected in the exhaled air.^14–16^ Larger particles with different sizes (between 1–50 µm) and compositions are exhaled more frequently during speech, laughter, or singing.^17–19^ In a recently published study from Singapore, it was shown that 85% of SARS-CoV-2 viruses were detected in the small size fraction of exhaled aerosols.^20^ The spread of viruses and bacteria via aerosols has already been investigated previously; e.g., in Mycobacterium tuberculosis, influenza viruses, and respiratory syncytial viruses (RSV),^7,21–23^ and aerosols are characterized as an important factor in the spread of related diseases. Aerosols are a vital transmission route for SARS-CoV-2 and play a major role in the viral spreading via asymptomatic individuals, contributing to the rapid spread of the SARS-CoV-2 pandemic.^7,8,10,13^

In outbreaks of other diseases, such as SARS and measles, previous studies have demonstrated that a small group (approximately 20%) of the primarily infected individuals was responsible for an estimated 80% of secondary infections;^9,24,25^ this is also true for to SARS-CoV-2.^26^ Recently, increased numbers of exhaled aerosol particles from a SARS-CoV-2-positive individual were reported on day eight of infection.^27^ In addition, Edwards et al. showed that in primates infected with SARS-CoV-2, there was a significant correlation between increasing levels of exhaled aerosols and the progression of pulmonary infection.^28^ According to these preliminary results, monitoring of exhaled aerosol particles (as a diagnostic tool) may be an important strategy in the mitigation of SARS-CoV-2 transmission. At present, most hygiene concepts are based on rapid antigen tests and PCR testing. Aerosol measurement may provide the advantage of direct on-site detection of infectivity within a few minutes; however, this test is unspecific because other airway infections may also be detectable.

If highly contagious individuals could be rapidly identified by aerosol measurement, and subsequently managed, a significant portion of new infections may be prevented. The aim of this prospective study was to investigate the difference in aerosol concentration and particle size between SARS-CoV-2 PCR-positive and -negative adults.

## Methods and Materials

### Study Design

We conducted a prospective observational cohort study to evaluate exhaled aerosol concentration and particle size in SARS-CoV-2 PCR-positive and -negative individuals at the Goethe University Hospital, Frankfurt, from February–June 2021. Eligible participants were adults (18–99 years) with a SARS-CoV-2 PCR test taken within 48 hours prior to aerosol measurement.

Before recruitment into the study, detailed verbal and written information was provided for all patients and controls; the aim and risks of the study were discussed in detail. Prior to the start of the study, written consent was obtained from all patients and controls. The study was approved by the Ethics Committee of the Goethe University Frankfurt (number 20-1001) and registered under the number ClinicalTrials.gov Identifier: NCT04739020. The study was sponsored by a grant of the Palas company (Karlsruhe Germany) detailed subsequently.

### Participants

Recruitment commenced on January 18th, 2021, and was completed on June 4th, 2021. In total, 288 adults were analyzed (64 subjects tested positive by a nasal or pharyngeal swab SARS-CoV-2 PCR and 224 healthy controls were SARS-CoV-2 PCR-negative). SARS-CoV-2 PCR-positive patients were recruited from the Division of Infectious Disease, Goethe-University Hospital, Frankfurt, Germany. Healthy controls were recruited from parents or caregivers of hospitalized children at the Department for Children and Adolescents, as well as volunteers. Volunteers were all fully vaccinated and asymptomatic. Caregivers and children were SARS-CoV-2 PCR-negative within 2 days prior to aerosol measurement and were quarantined until measurement was conducted; hospitalized children were admitted for diagnoses other than SARS-CoV-2 infection. SARS-CoV-2 PCR test was obtained when clinically feasible, in patients admitted with a positive SARS-CoV-2 PCR test before admission. Subjects were excluded from study entry if unable to participate in aerosol measurement, perform spirometry, or understand the extend and consequences of the study.

### Study Procedures

#### Clinical and Medical History

The electronic chart and International Classification of Disease (ICD) were used for diagnosis, estimation of BMI, oxygen supplementation, and to cluster risk factors including obesity, diabetes, hypertension, and chronic heart, respiratory, and kidney disease.

Before measurement of aerosols, all participants (SARS-CoV-2 PCR-positive patients and healthy controls) were questioned about the presence of typical SARS-CoV-2 symptoms. The typical SARS-CoV-2 symptoms included the presence of fever, cough or dry cough, shortness of breath, loss of taste or smell, sore throat, muscle pain, diarrhea, and vomiting.

#### Aerosol measurement

Exhaled particles were counted and sized by an aerosol spectrometer (Resp-Aer-Meter, Palas GmbH, Karlsruhe, Germany), specifically designed to detect airborne exhaled particles in the size range of 0.15–5.0 μm with very high sizing resolution (16 channels/decade). The optical sensor utilized a polychromatic light source to create a defined optical measurement volume, with every particle travelling through generating a scattered light pulse. The size and quantity of particles were determined from the number and intensity of the scattered light pulses.

The instrument compromised a heated hose section upstream of the measurement cell to avoid condensation effects and enable evaporation of larger droplets. The temperature and relative humidity in the sampled air was also measured. Exhaled breath from subjects was collected via mouthpiece, connected to a t-adapter with HEPA filter and connection port to the Resp-Aer-Meter via a hose. To ensure effective hygiene, sterile sampling kits were used for each measurement. Participants performed normal tidal breathing through the mouthpiece while the nose was closed via a nose clip. In the first minute of tidal breathing, a sharp drop of particle concentration was detected due to inhalation of clean air via the HEPA filter. This is the washout effect, during which the ambient aerosol still present in the lungs is washed out. After a few breaths, a baseline concentration of particles generated and exhaled from the lungs was determined. Subsequently, the measurement (lasting 1–1.5 minutes) was taken to establish the quantity of particles emitted from the lungs. The results of the test were directly displayed as a graphical curve (Supplemental eFig. 1), enabling calculation of the mean exhaled particle count per liter, particle size distribution, and mean particle size (in µm).

#### Spirometry

Spirometry was performed according to the recommendations of the American Thoracic Society (ATS) and the European Respiratory Society (ERS)^36^ by a hand-held device (Asthma Monitor® AM; VIASYS Healthcare GmbH, Höchberg, Germany). Measurements of Peak Expiratory Flow (PEF) and Forced Expiratory Volume in the first second (FEV1) were obtained.

### Outcomes

The primary outcome of this study was the measurement of aerosol particle concentration in SARS-CoV-2 PCR-positive and -negative subjects, and the distinction between positive and negative subjects via aerosol measurement. Secondary outcome measures comprised the correlation of aerosol concentration to SARS-CoV-2 PCR results, change in aerosol concentration due to confounders (such as age, sex, lung function, height, weight, BMI, and smoking status), and the correlation between clinical symptoms and aerosol measurements in SARS-CoV-2 PCR-positive patients.

### Statistical analysis

GraphPad Prism 5.01 (GraphPad Software, Inc.) and R 4.0.4 were used for statistical analysis. The values were presented as median and range for numeric data and as percentage for count data.

The Wilcoxon-Mann-Whitney U-Test and Fisher’s exact test were used to test for group differences in numeric and count data, respectively. A p-value of less than 0.05 was considered statistically significant.

In addition, the sensitivity and specificity of the aerosol measurement was evaluated using Receiver Operating Characteristic (ROC) analysis and the correlation between Ct values and aerosol measurement was calculated via Spearman correlation.

## Results

### Patient Characteristics

SARS-CoV-2 PCR-positive patients were older (median 53 years vs. 41 years; p<0.0001), predominantly male (68.8% vs. 30.4%; p<0.0001), and with higher BMI (median 28.4kg/m^2^ vs. 25.6kg/m^2^; p<0.001) when compared with the healthy control group. Smoking status did not differ between groups (p=0.678). Detailed characteristics of all groups are presented in Table 1.

**Table 1:** Characteristics of PCR SARS-CoV-2 PCR-positive and -negative patients.

|  | **Respiratory**<br>**failure**<br>**(n=12)** | **Pneumonia**<br>**(n=34)** | **Immuno-**<br>**compromised**<br>**(n=18)** | **SARS-CoV-**<br>**2 PCR-**<br>**positive**<br>**(n=64)** | **SARS-CoV-2**<br>**PCR-negative**<br>**(n=224)** | **p-value** |
| --- | --- | --- | --- | --- | --- | --- |
| **Sex** |  |  |  |  |  |  |
| female | 2 (16.7%) | 14 (41.2%) | 4 (22.2%) | 20 (31.3%) | 156 (69.6%) | <0.0001 |
| male | 10 (83.3%) | 20 (58.8%) | 14 (77.8%) | 44 (68.8%) | 68 (30.4%) |  |
| **Smoking status** |  |  |  |  |  |  |
| Non-smoker | 11 (91.7%) | 27 (81.8%) | 12 (66.7%) | 50 (79.4%) | 173 (77.2%) | 0.678 |
| Smoker | 1 (8.3%) | 6 (18.2%) | 6 (33.3%) | 13 (20.6%) | 51 (22.8%) |  |
| **BMI (kg/m<sup>2</sup>)** |  |  |  |  |  |  |
| Median | 26.7 | 29.2 | 26.4 | 28.4 | 25.7 | <0.001 |
| Range | 24.2–44.4 | 16.6–40.3 | 18.8–41.3 | 16.6–44.4 | 17.0–45.0 |  |
| **Age (years)** |  |  |  |  |  |  |
| Median | 57 | 58 | 51 | 53 | 41 | <0.0001 |
| Range | 34–85 | 28–87 | 24–78 | 24–87 | 18–80 |  |
| **FEV1 (%pred.)** |  |  |  |  |  |  |
| Median | 72.2 | 48.2 | 73.9 | 58.1 | 86.7 | <0.0001 |
| Range | 28.6–97.1 | 16.5–82.9 | 49.1–107.7 | 16.5–107.7 | 55.9–149.3 |  |
| **CT value** |  |  |  |  |  |  |
| Median | 28.4 | 27.5 | 21.9 | 26.8 | >40.0 |  |
| Range | 20.4–36.1 | 15.8–37.4 | 16.4–37.5 | 15.8–37.5 |  |  |
| **Comorbidities** |  |  |  |  |  |  |
| Obesity | 5 (41.7%) | 14 (41.2%) | 3 (16.7%) | 22 (34.4%) | 44 (19.6%) | 0.0066 |
| Hypertension | 2 (16.7%) | 13 (38.2%) | 7 (38.9%) | 22 (34.4%) | 13 (6.4%) | <0.0001 |
| Diabetes | 4 (33.3%) | 8 (23.5%) | 2 (11.1%) | 14 (21.9%) | 3 (1.5%) | <0.0001 |
| Respiratory<br>Disease | 1 (8.3%) | 8 (23.5%) | 0 (0.0%) | 9 (14.1%) | 24 (10.7%) | 0.460 |
| Cardiac Disease | 1 (8.3%) | 7 (20.6%) | 4 (22.2%) | 12 (18.8%) | 0 (0.0%) | <0.0001 |
| Kidney Disease | 0 (0.0%) | 9 (26.5%) | 9 (50.0%) | 18 (28.1%) | 0 (0.0%) | <0.0001 |

SARS-CoV-2 PCR-positive group is displayed as whole collective and divided in three clinical categories (respiratory failure, pneumonia, immunocompromised). p-Values for differences in SARS-CoV-2 PCR-positive and -negative participants are derived from Wilcoxon-Mann-Whitney U-Test for numeric data and from Fisher's exact test for count data.

### Clinical and Medical History

Of 64 hospitalized SARS-CoV-2 PCR-positive patients, 71.9% (46/64) were diagnosed with acute respiratory failure and/or pneumonia associated with SARS-CoV-2 infection: 12 patients were diagnosed with respiratory failure and 34 with COVID-19 pneumonia. In total, 28.1% (18/64) of patients had moderate symptoms and were considered immunocompromised with precautionary hospital admissions when found to be SARS-CoV-2 PCR-positive. A majority (15/64) presented with an underlying oncological condition, but 3/64 were admitted with other pre-existing medical conditions.

In the SARS-CoV-2 PCR-positive group, 76.6% (49/64) of patients had pre-existing medical conditions (Table 1). In the control group, subjects reported the following pre-existing medical conditions: 3.6% (8/224) had asthma, 4.0% (9/224) had allergies, 7.9% (16/224) had hypothyroidism, 6.44% (13/224) had arterial hypertonia, and 1.5% (3/224) had diabetes mellitus.

The symptoms recorded in the SARS-CoV-2 PCR-positive and -negative subjects are presented in Table 2.

**Table 2:** Clinical symptoms according to questionnaire.

| **Symptoms** | **Respiratory**<br>**Failure**<br>**(n=12)** | **Pneumonia**<br>**(n=34)** | **Immuno-**<br>**compromised**<br>**(n=18)** | **SARS-**<br>**CoV-2**<br>**PCR-pos.**<br>**(n=64)** | **SARS-**<br>**CoV-2**<br>**PCR-neg.**<br>**(n=224)** | **p-value** |
| --- | --- | --- | --- | --- | --- | --- |
| **Fever** | 4 (33.3%) | 10 (29.4%) | 3 (16.7%) | 17 (26.6%) | 0 (0.0%) | <0.0001 |
| **Cough** | 7 (58.3%) | 25 (73.5%) | 2 (11.1%) | 34 (53.1%) | 4 (1.8%) | <0.0001 |
| **Dyspnea** | 8 (66.7%) | 18 (52.9%) | 0 (0.0%) | 26 (40.6%) | 1 (0.5%) | <0.0001 |
| **Loss of**<br>**taste/smell** | 3 (25.0%) | 6 (17.6%) | 1 (5.6%) | 10 (15.6%) | 0 (0.0%) | <0.0001 |
| **Sore**<br>**throat** | 4 (33.3%) | 4 (11.8%) | 0 (0.0%) | 8 (12.5%) | 0 (0.0%) | <0.0001 |
| **Myalgia** | 4 (33.3%) | 4 (11.8%) | 0 (0.0%) | 8 (12.5%) | 0 (0.0%) | <0.0001 |
| **Diarrhea** | 1 (8.3%) | 4 (11.8%) | 1 (5.6%) | 6 (9.4%) | 1 (0.5%) | <0.0001 |
| **Vomiting** | 0 (0.0%) | 2 (5.9%) | 0 (0.0%) | 2 (3.1%) | 0 (0.0%) | 0.064 |

Symptoms were recorded at time of aerosol measurement. SARS-CoV-2 PCR-positive group is displayed as whole collective and divided in three clinical categories (respiratory failure, pneumonia, immunocompromised). p-Values for differences in SARS-CoV-2 PCR-positive and -negative participants are derived from Fisher's exact test.

### Aerosol Measurements

#### Aerosol Concentration

The median exhaled particle count was highly significantly elevated in SARS-CoV-2 PCR-positive patients (1490.5/L [46.0–34,772.0/L]) compared with healthy controls (252.0/L [0.0–882.0/L]; p<0.0001, Fig. 1). This significant difference between SARS-CoV-2 PCR-positive and negative patients was confirmed by an age-matched control group (Supplemental eFig. 2)

**Figure 1:**
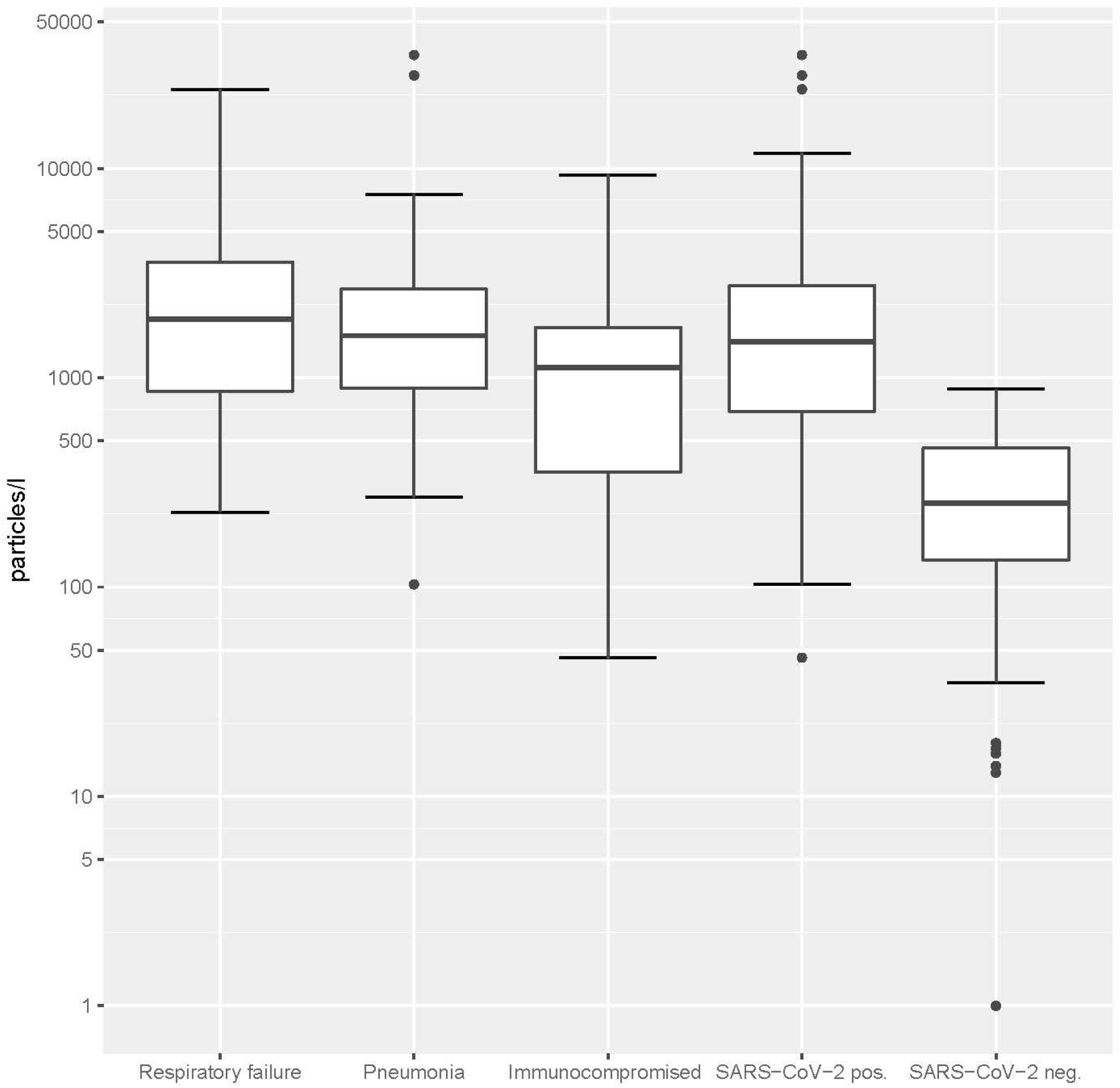
Aerosol particle counts in SARS-CoV-2 PCR-positive and -negative patients. SARS-CoV-2 PCR-positive patients are displayed as a collective and divided in three clinical subgroups (respiratory failure, pneumonia, immunocompromised).

Exhaled particle counts >5,000/L were considered very high, with the expectation of elevated contagiousness in the viral infection setting. In the SARS-CoV-2 PCR-negative group, no subjects (n=0/224) reported very high exhaled particle counts; in the SARS-CoV-2 PCR-positive group, 15.6% (n=10/64) showed high counts and were responsible for 64.8% of all exhaled particle counts in the group. Moreover, the 15.6%, equating to 3.5% of all patients (n=10/288), was responsible for 51.2% of all exhaled particles.

In addition, there was a significant, negative correlation for exhaled particle count and Ct value (Spearman correlation, r: -0.4926; p<0.0001).

There were no significant differences in aerosol concentration due to sex, BMI, or smoking status (Supplemental eFig. 3); however, there was a slight increase of aerosol concentration with greater age.

When considering the SARS-CoV-2 PCR-positive group only, there was a slight difference in median exhaled particle counts across the three subgroups respiratory failure (1953.0/L [228.0–23,861.0/L]), pneumonia (1586.5/L [103.0–34,772.0/L]), and immunocompromised patients (1122.0/L [46.0–9,319.0/L]; p=0.19, Fig. 1).

#### Aerosol Particle Size

Regarding the particle size distribution, the available size channels (in total, 14 size channels from 0.15–5.0 µm) were analyzed in across three size bands: <0.3 µm, 0.3– 0.5 µm, and >0.5–5.0 µm. For both groups, the majority of the aerosols (>90% in the SARS-CoV-2 PCR-positive group and >78% in the -negative group) were found in the smallest range (<0.3 µm). Especially for the positive group, increases in total aerosol concentration were dominated by increases in particles ≤0.3 µm.

#### ROC Analysis of Aerosol Concentration

In order to analyze the accuracy of the exhaled particle count as a test to detect SARS-CoV-2 PCR-positive infection, a ROC analysis was conducted (Fig. 2). At an exhaled particle count cut-off value of 596/L, the sensitivity of the test was 79.7%, and specificity was 85.7%, with an area under the curve (AUC) of 0.8918.

**Figure 2:**
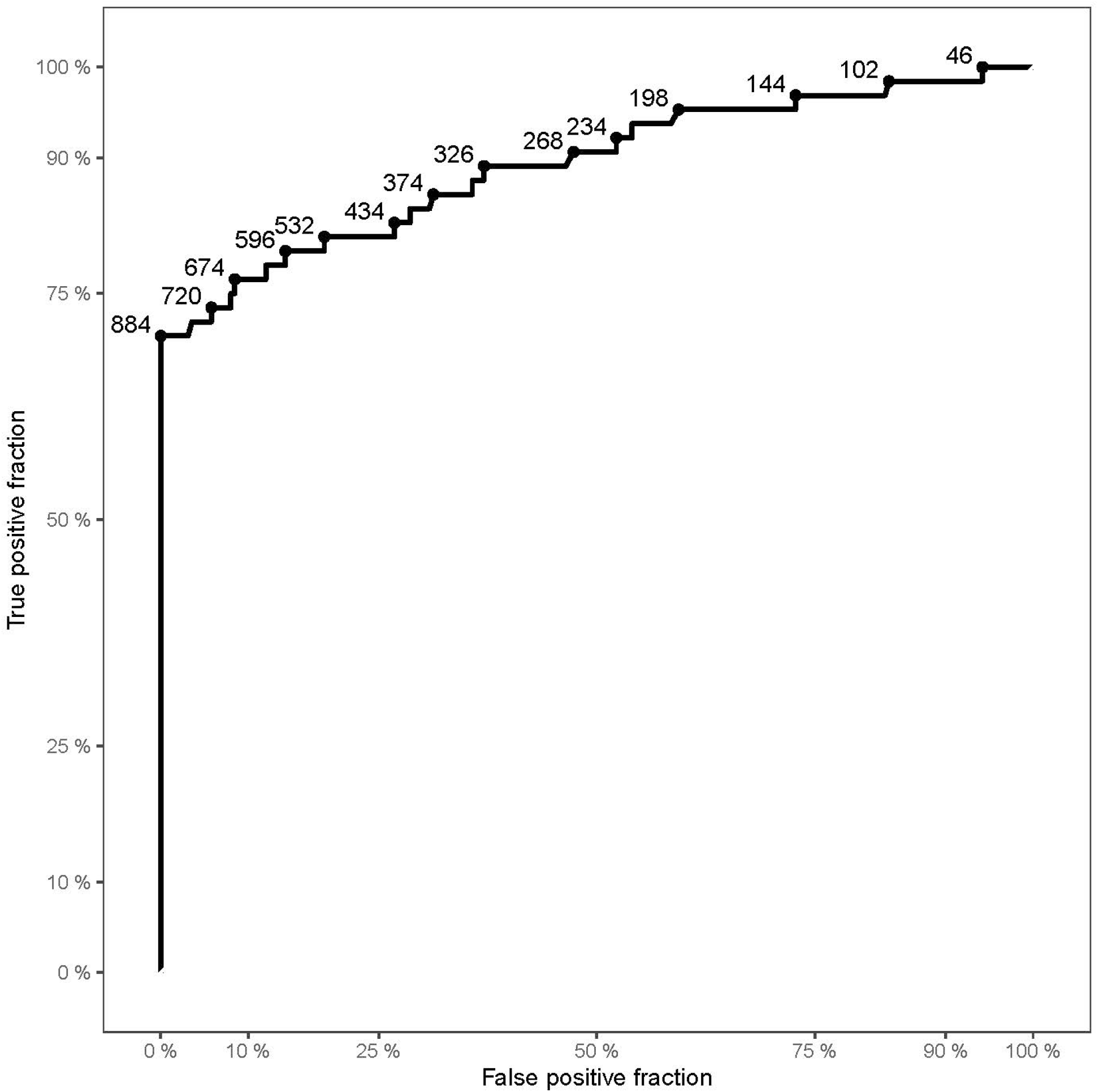
ROC curve of the dataset. Displaying sensitivity (true positive fraction) in the y-axis and 1-specificity (false positive fraction) on the x-axis. Points on the curve show examples of cut-off values (aerosol particles per liter) with corresponding sensitivity and specificity.

### Spirometry

To account for age, weight, and sex during FEV1 analysis, the FEV1%pred (FEV1 in percentage of the predicted value) was calculated. The median FEV1%pred in the SARS-CoV-2 PCR-positive and -negative groups was 58.1% and 86.7% (p<0.0001), respectively.

In addition, there was a significant difference of FEV1%pred between the patient (respiratory failure, pneumonia, and immunocompromised) and healthy control groups (Table 1).

## Discussion

Current research suggests that SARS-CoV-2 infection is spread primarily through exhalation of droplets and aerosols containing viable virus particles, which may linger in the air and survive for several hours.^6,17,30^ Despite inter-individual differences, SARS-CoV-2 PCR-positive patients produced significantly increased exhaled particle counts compared with healthy controls. In addition, greater exhaled particle counts may be associated with more severe infection and higher infectivity.

Whereas Edwards et al. reported a significant correlation between exhaled particle counts and BMI,^28^ no such correlation was observed within either the SARS-CoV-2 PCR-positive or -negative groups in this study. In addition, no correlation in particle counts was found for smoking status or sex. However, similar to Edwards et al.,^28^ there was a correlation between exhaled particle counts and age, with greater counts observed with increasing age. It should be considered that Edwards et al. utilized a different aerosol detector able to detect particles in a size range >0.3 µm. The current study suggests that the majority of exhaled particles are <0.3µm; thus, these data are not completely comparable.

Lästard et al.^31^ and Almstrand et al.^32^ determined that small, exhaled aerosol particles consist mainly of surfactant. Edwards et al. found that an increase of surfactant in the lungs generates significantly more exhaled aerosol particles.^24^ It is recognized that the alveolar epithelial type 2 (AT2) cells in the lungs are responsible for surfactant production and also have ACE2 receptors (which are the port of entry for SARS-CoV-2 viruses into the human cells). Therefore, it seems likely that SARS-CoV-2 viruses are replicated in AT2 cells; these cells consecutively may release more surfactant into the airways, which may be responsible for a significant increase of small aerosol particles by reopening of collapsed airways. Within these aerosol particles, SARS-CoV-2 viruses are transported from the lungs and can be inhaled by others to spread viral infection.

More than 60 years ago, Wells et al. reported that small aerosol particles (<5 µm) may remain airborne indefinitely while indoors.^33^ Other authors have found that large droplets remain in the upper respiratory tract, whereas smaller particles travel down the respiratory tract to the bronchi.^21,22,33-35^ In this study, exhaled aerosols from SARS-CoV-2 PCR-positive patients produced a greater number of small particles, as well as significantly lower mean particle sizes, compared with healthy controls.

Recent studies have demonstrated that increases in exhaled particle concentration with SARS-CoV-2 positive primates are dominated by very small particles, which might only be visible with a lower detection limit below 0.3 µm.^28^ Thus, measurement with a lower detection limit may provide greater accuracy for detecting exhaled particles, particularly in the size ranges that are crucial for transmission of SARS-CoV-2.

Jones et al. highlighted a large-sample analysis of RT-PCR results, which showed that a small subset of subjects (9%) had a very high viral load and were thus considered highly infectious.^36^ The current study demonstrated that a very small group (3.5% of all participants) was responsible for over 50% of all exhaled aerosols. Furthermore, in the SARS-CoV-2 PCR-positive group, 15.6% of patients were responsible for almost 70% the of exhaled aerosols.

To assess the validity of the aerosol measurement as a tool to test for SARS-CoV-2 infected patients, a ROC analysis was conducted and demonstrated good validity (AUC of 0.89). Our analysis suggests that aerosol particle measurements alone are not sufficiently sensitive (79.7%) nor specific (85.7%) to diagnose a SARS-CoV-2 infection, but can provide complementary diagnostic information when used in combination with SARS-CoV-2 PCR tests, or rapid antigen testing. In addition, as patients with high aerosol counts appear to suffer from more severe disease, elevated aerosol counts may be associated with increased severity and contagiousness; future studies are required to confirm this hypothesis. In addition, depending on selection of cut-off values, aerosol measurement could provide potential utility as a screening tool in select individuals who require further PCR testing.

Our study has several limitations, including that the SARS-CoV-2 PCR and aerosol particle measurements were not performed simultaneously. A timeframe from PCR test to aerosol measurement of 72 hours was accepted for all patients; this may affect the results, as other studies have found peak viral loads around day four of infection, which might be present 1-3 days before the onset of symptoms and followed by a steady decline in viral load.^36^ In addition, only hospitalized SARS-CoV-2 positive patients were included. And it seems reasonable that aerosol particle counts may be greater in patients with severe disease, reflecting a certain level of lung framework damage due to this viral infection. This might explain the lower exhaled particle counts reported in the immunocompromised sub-group, when compared with the pneumonia and respiratory failure groups. Further studies should assess whether similar quantities of aerosol particles are produced by asymptomatic individuals and patients with mild infection. The utility of this approach as a diagnostic tool for patients at earlier stages of infection, a critical time for diagnosis, is not addressed by this work and will be the focus of further research. Moreover, the duration of elevated particle counts is unclear in SARS-CoV-2 PCR-positive patients. This may only be verified by longitudinal measurements of aerosol particles and serial SARS-CoV-2 PCR measurements. In addition, this study allows no statement concerning differences in viral load^36^ and consecutive aerosol shedding by virus variants, although these were previously observed clinically. Lastly, only aerosols across a particular size range were measured. While current research suggests that small particles may play a critical role in aerosol transmission, future studies investigating different aerosol size ranges should be performed to verify this hypothesis.

In conclusion, the concentration of exhaled aerosols particles was significantly different between SARS-CoV-2 PCR-positive and -negative individuals. Because the origin of these aerosol particles are the bases of the lung, alveolar epithelial cells type 2 may produce more surfactant when infected by viruses, generating more small droplets to carry the virus out of the lung. A better understanding of respiratory aerosol generation may lead to improved control of SARS-CoV-2 transmission. In the future, portable devices for aerosol measurement may be a valuable tool to detect potentially contagious individuals with a non-invasive breath test.

## Data Availability

Anonymized participant data will be made available upon requests directed to the corresponding author. Proposals will be reviewed and approved by investigator and collaborators on the basis of scientific merit. After approval of a proposal, data can be shared through a secure online platform after signing a data access agreement. All data will be made available for a minimum of five years from the end of the trial.

## Role of funding source

This work was supported by Palas GmbH, Partikel-und Lasermesstechnik, which provided aerosol measurement devices, as well necessary equipment, and limited sponsorship for conduction of the study. Additional resources were provided by the University Hospital Frankfurt, Goethe University.

Palas GmbH reviewed the study data and final manuscript before submission, but the authors retained editorial control. Palas GmbH also provided funding for a medical writer to assist in manuscript preparation.

All authors had full access to all data in the study and had final responsibility for the decision to submit for publication.

## Acknowledgements

We would like to thank Dr Allan Johnson of Medical Writing Limited for his assistance in preparation and editing of this manuscript. We also thank the volunteers who participated in this study.

## Contributors

DG, HD, MH, RS, MW, and SZ conceived and designed the trial and SZ was the principal investigator. DG, HD, LH, TL and YK, collected trial data. DG, HD, TL, LH, YK, AG, GS and FW were involved with data curation, and DG, MH, and FK were responsible for the formal data analysis. DG, MH, TL, LH, GS and SZ verified the underlying data supporting the findings of this manuscript. All authors had full access to the full data set in this study and critically reviewed and approved the final version.

