## Supplemental data for "Aerosol measurement identifies SARS-CoV 2 PCR positive adults compared with healthy controls"

Methods

eFigure 1: Aerosol Resp-Aer-Meter (A) structure and (B) function, as well as (C) sample measurement.

eFigure 2: Aerosol particle counts in age-matched SARS-CoV-2 PCR-positive and -negative patients.

eFigure 3: Aerosol particle counts in SARS-CoV-2 PCR-positive and -negative patients divided into subgroups.

**Methods**

SARS-CoV-2 PCR

Repeatedly performed combined nasal and throat PCR swab tests were confirmed before and during admission in all subjects. Semiquantitative real-time PCR (RT-PCR) was assessed predominantly by commercial PCR test. Test results were reported as ‘negative’ or ‘positive’; in the case of a positive result, the cycle threshold (Ct) value was reported. In all participants, a SARS CoV-2 PCR was obtained before enrollment.

**eFigure 1: Aerosol Resp-Aer-Meter (A) structure and (B) function, as well as (C) sample measurement.**

**A. Aerosol Resp-Aer-Meter structure**

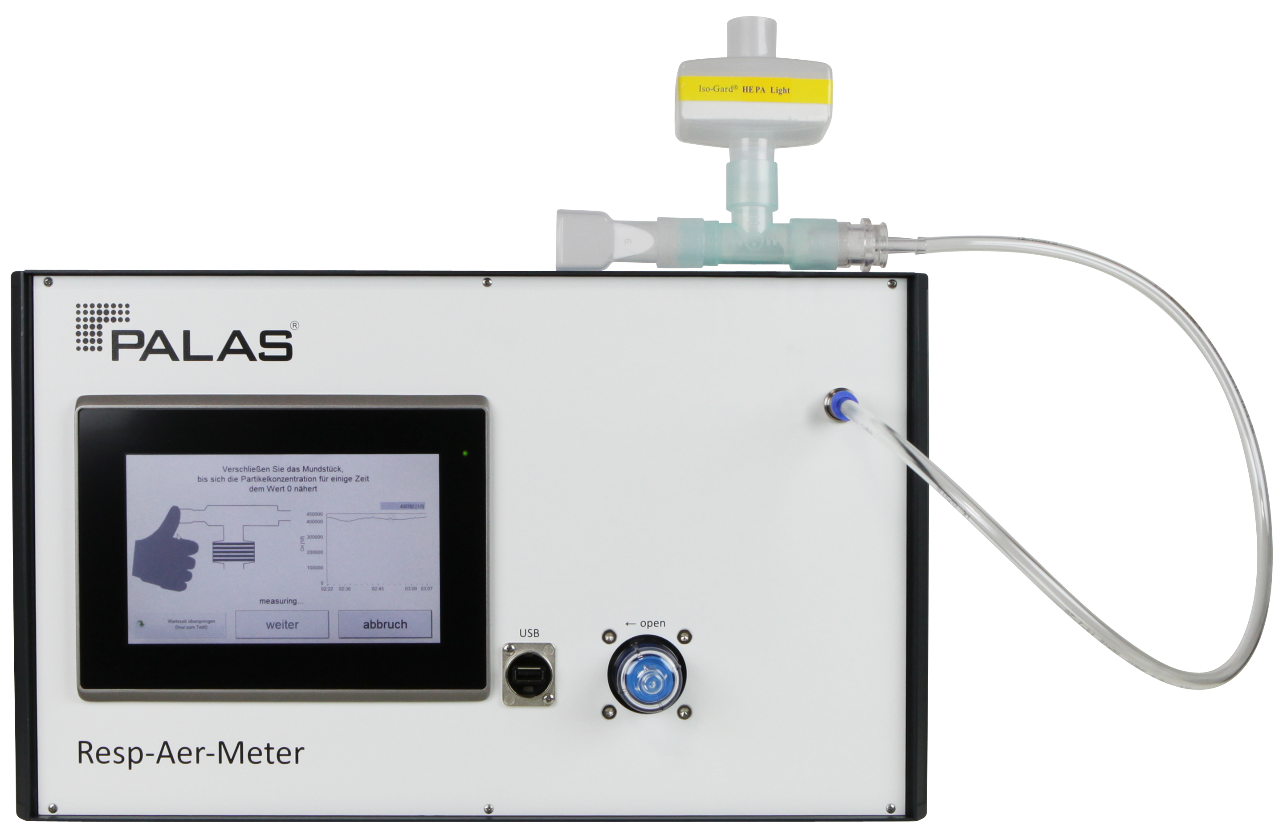

**B. Aerosol Resp-Aer-Meter function**

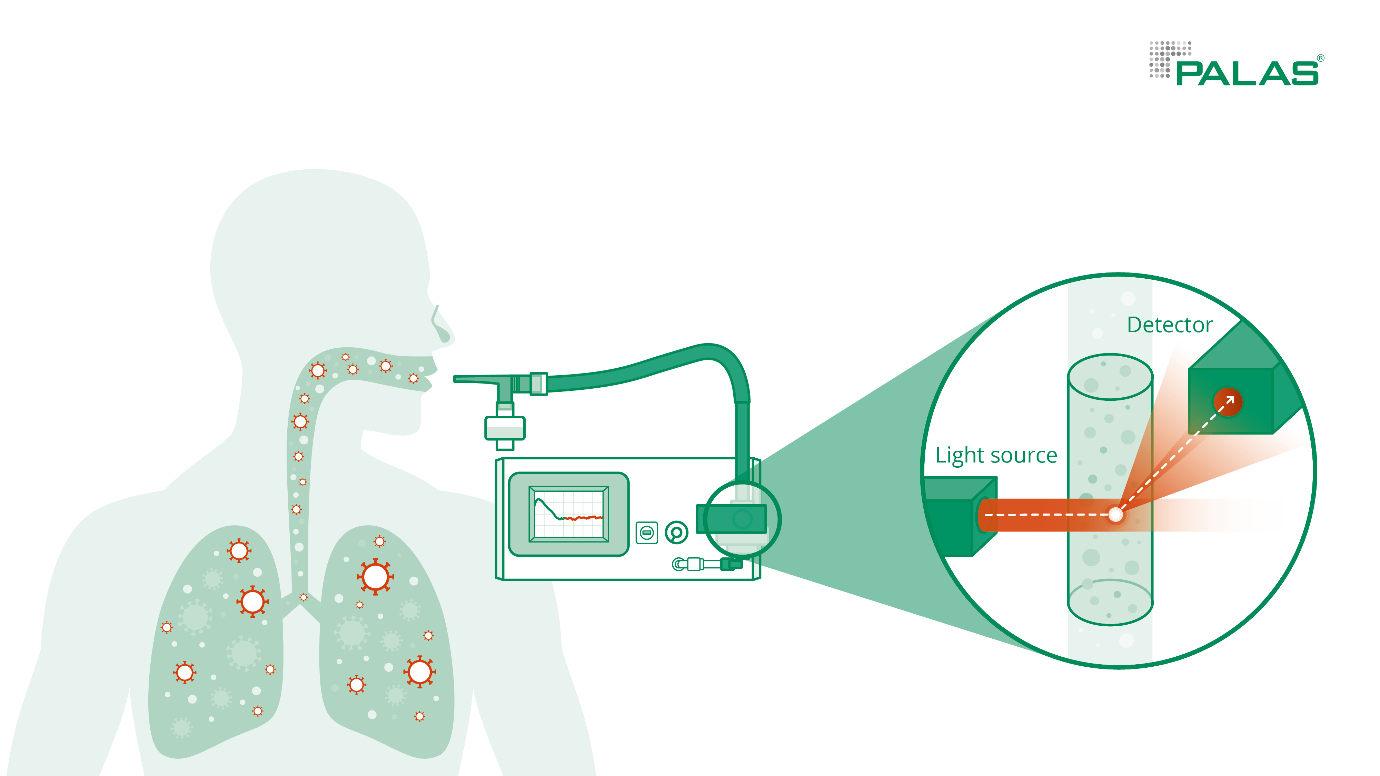

**C. Aerosol Resp-Aer-Meter sample measurement**

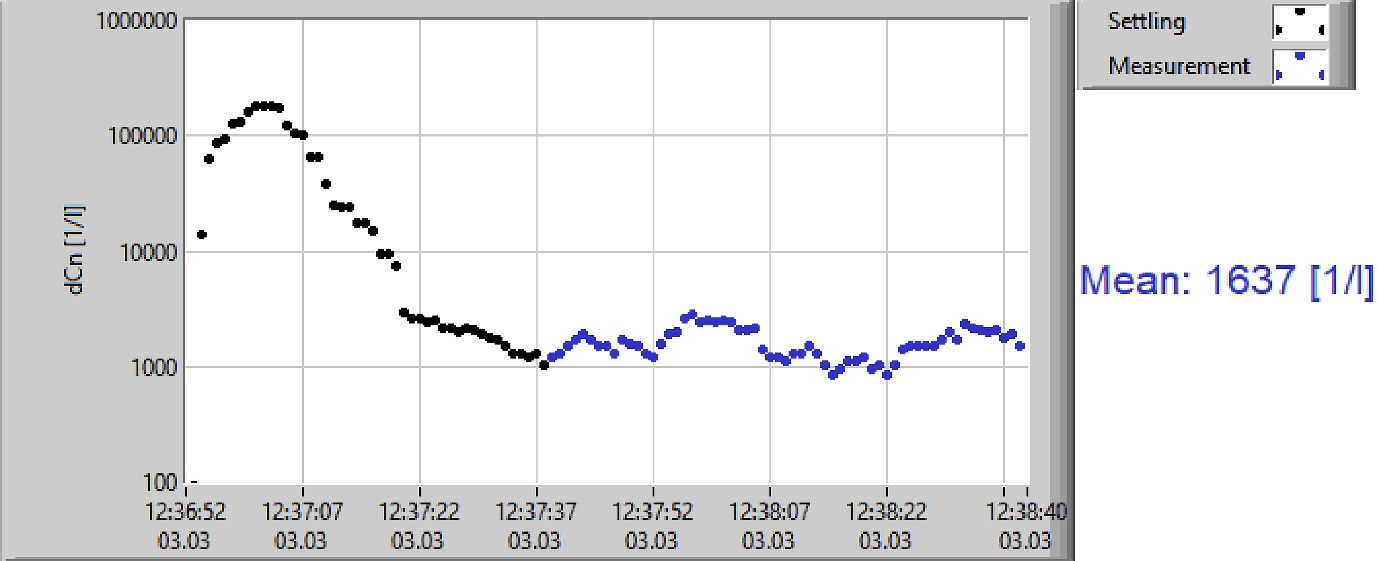

**eFigure 2: Aerosol particle counts in age-matched SARS-CoV-2 PCR-positive and -negative patients.**

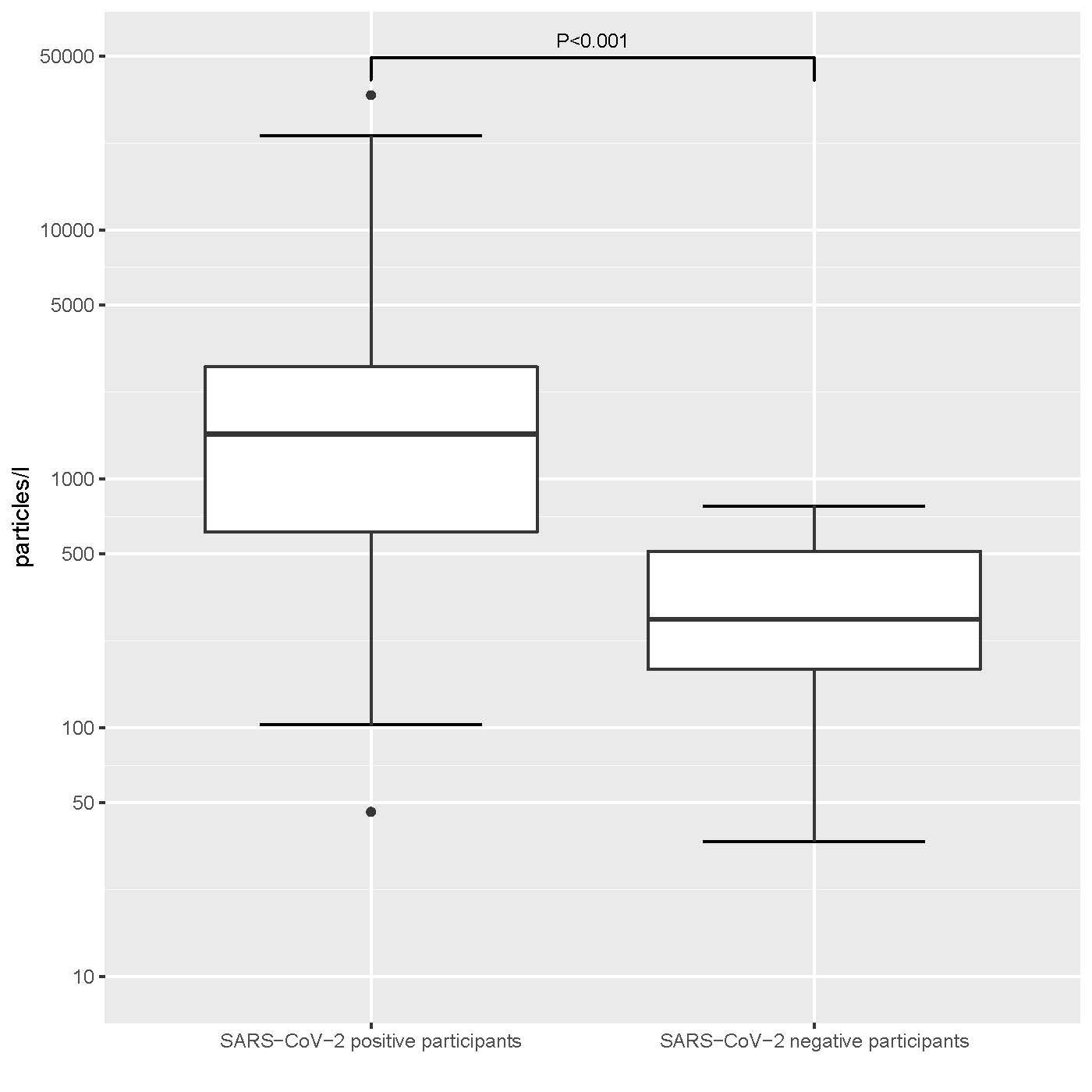

**eFigure 3: Aerosol particle counts in SARS-CoV-2 PCR-positive and -negative patients divided into subgroups.**

**A. Divided into female and male patients**

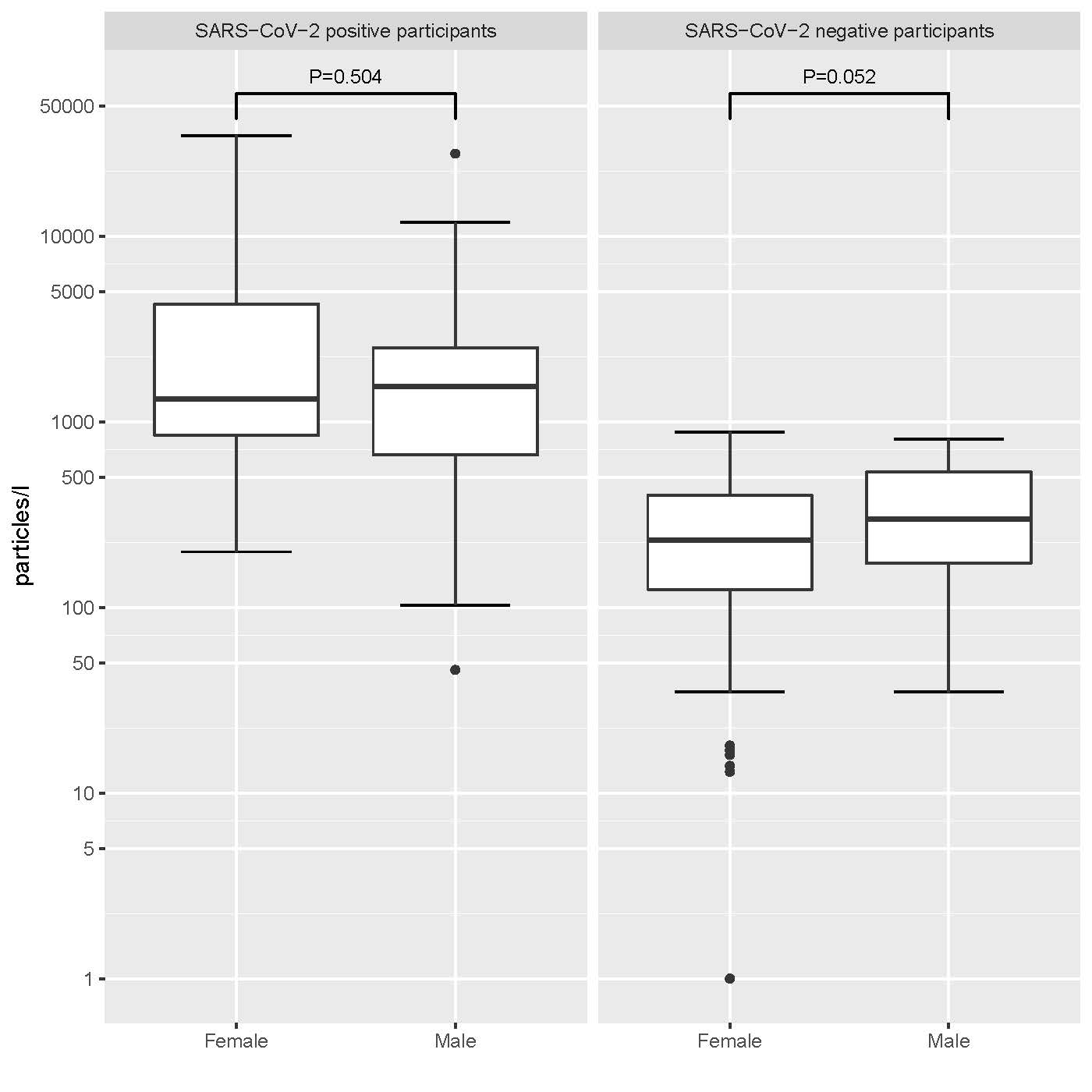

**B. Divided in groups of BMI ≤30kg/m^2^ and >30kg/m^2^**

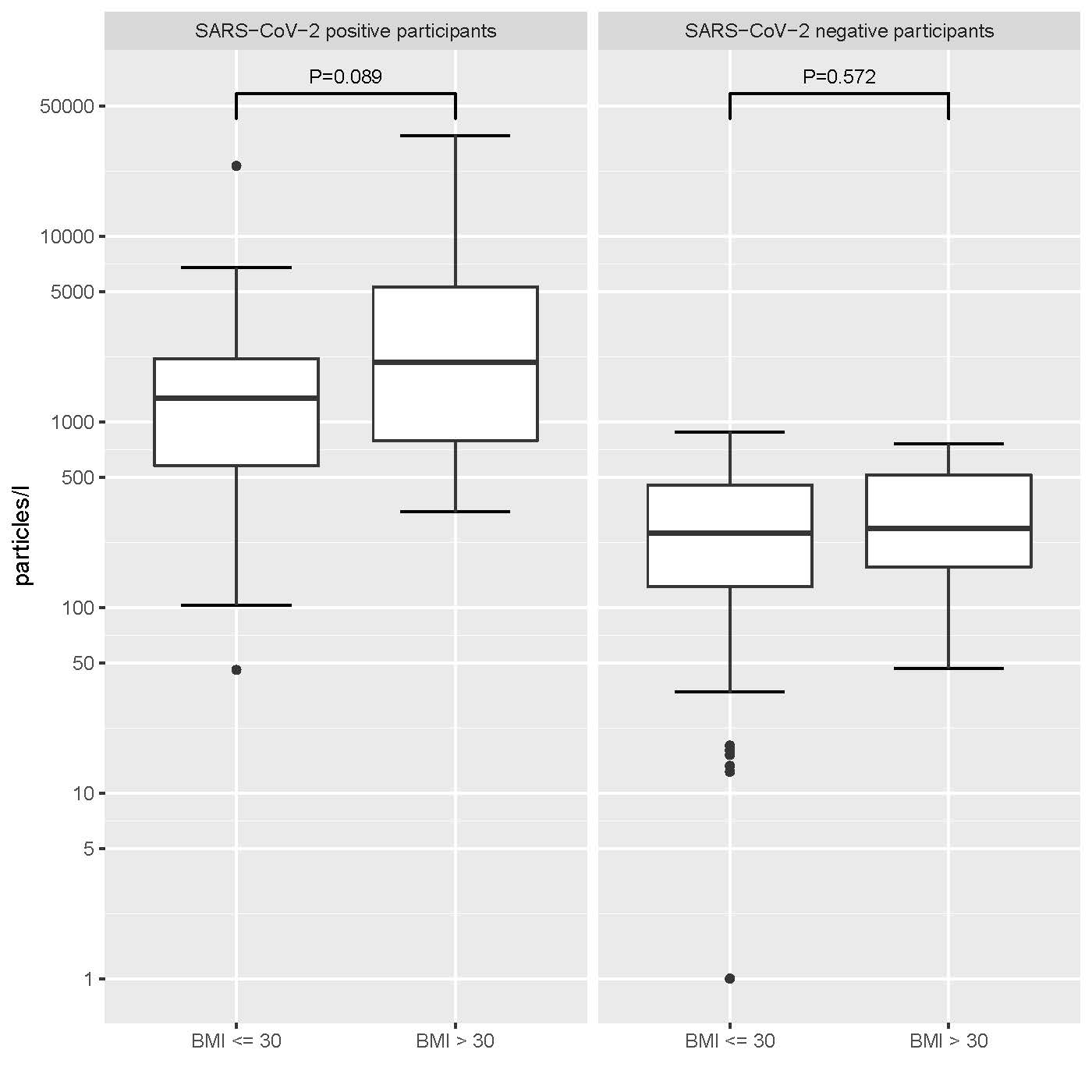

**C. Divided into non-smoker and smoker.**

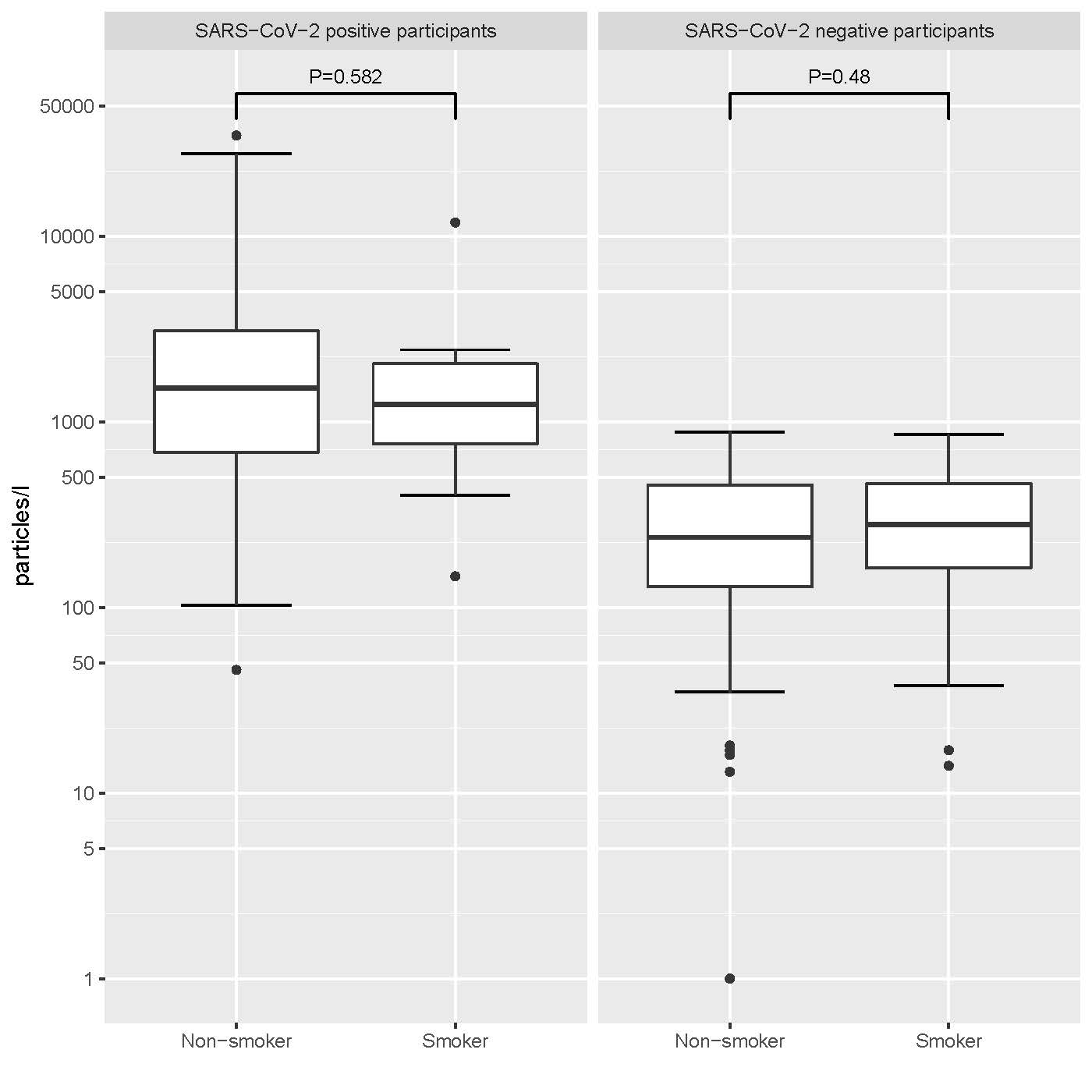
